# Over-expressed KRT8 in the cytotrophoblast shell promotes the development of preeclampsia

**DOI:** 10.1101/2025.01.06.25320086

**Authors:** Miaomiao Chen, Xiaohong Yang, Xiangyi Chen, Hen Yin, Junbo Wu, Jing Peng, Menghan Sha, Chunyan Liu, Qianwen Dai, Kai Zhao, Yun Zhao

## Abstract

Preeclampsia is a “placenta-derived disorder” characterized disease, which remains a major unaddressed public health problem. The differentiation and development of cytotrophoblasts and extravillous trophoblasts are two crucial processes, which are tightly regulated. And any abnormal regulation can lead to the occurrence of pregnancy-related diseases such as preeclampsia. In this article, we performed Spatial transcriptomics on tissues of PE and Normal, and found a large amount of extravillous trophoblasts (EVTs) were accumulated in the cluster 8 (cytotrophoblast shell) of PE_decidua, and Trajectory analysis revealed KRT8 was over-expressed in the cytotrophoblast shell of PE_decidua. The accumulation of EVTs caused by the increase of KRT8 promotes the development of preeclampsia.

## Introduction

Throughout the pregnancy, a successful pregnancy largely depends on its placenta, which is located at the mother-fetus interface.^[1]^ In the placental villi, villous cytotrophoblast (CTB) fuse to form the overlying syncytiotrophoblast (SCT) layer that is in contact with maternal blood in the inter villous space. And then cytotrophoblast cell columns (CCCs) break through the SCT and arise from the CTBs.^[2, 3]^ After that, extravillous trophoblasts (EVTs) which emerge from the cytotrophoblast shell, begin to differentiate in cell columns but invasive EVTs emerge only when the anchoring villi attach to the maternal decidua.^[4]^ Interstitial trophoblast cells (iEVTs) subsequently fuses into placental bed giant cells (GCs) around the deciduation-myometrium boundary, usually invading only the inner third of the myometrium1^[5]^ Subsequently, endovascular trophoblast cells (eEVTs) form a plug close to the cytotrophoblast shell where the arteries terminate and then eEVTs replace the endothelium.^[6, 7]^

Preeclampsia (PE) is a “placenta-derived disorder” characterized by new-onset high blood pressure after 20 weeks of pregnancy with multisystem damage, which often leads to progressive multiorgan dysfunction. It affects approximately 2-4% of pregnant women worldwide and causes approximately 46,000 maternal deaths and 500,000 fetal and neonatal deaths each year.^[8]^ The pathogenesis of preeclampsia is still not fully understood. Impaired uterine spiral artery remodeling (SAR) and placental implantation can be found in preeclampsia.^[9, 10]^ However, to date, the molecular mechanisms connecting dysregulated uterine immune cells and impaired tissue remodeling in preeclampsia remain to be defined. One of the challenges has been to define the landscape of individual cell populations at the maternal–fetal interface and to understand how those cells function spatiotemporally under pathophysiological conditions.

The emergence of spatial transcriptomics (ST) makes it possible to comprehensively delineate the landscape of the PE placenta and elucidate of the pathogenesis of PE. In February 2023, the Roser Vento-Tormo team^[4]^ described the complete trajectory of trophoblastic invasion during early trimester through single-nuclei RNA sequencing (snRNA-seq), combined snRNA-seq and single-nuclei assay for transposase-accessible chromatin with sequencing (snATAC-seq) and ST using Visium. On the other hand, the establishment of placental perfusion is inseparable from spiral artery remodeling, and many diseases in pregnancy are characterized by inadequate spiral artery remodeling. In July, the authors divided spiral artery remodeling into five stages to study the invasion process of EVTs that we have been concerned about.^[7]^ Besides, the authors also provided image-based and statistical answers to some scientific questions on the maternal-fetal interface, including the dynamics of the immune micro-environment around the spiral artery and the decidual immune micro-environment during pregnancy which can help us to further understand EVT cells in detail^[7]^. ST technologies have emerged as tools that can not only capture the spatial context of RNA molecules but also complement Single-cell transcriptomics (scRNA-seq) for biological discoveries.^[11-13]^ In this article, we applied the ST technologies to understand the pathophysiology of PE.

## Materials and methods

### Sample collection

The whole layer of the placenta (from decidua to villi, 1^*^1^*^2cm^3^) was harvested from a distance of 2 cm from the center of the umbilical cord, and the tissue was divided into two parts(1^*^1^*^1cm^3^), respectively named as decidua (including the decidual part), and villus (the remaining part). All tissues and serum were collected from pregnant women who underwent caesarean delivery at Maternal and Child Health Hospital of Hubei Province, Affiliated Hospital of Tongji Medical College, Huazhong University of Science and Technology at July 2024. The participants provided a signed informed consent. Patients with pregnancy complications and complications like diabetes, cardiovascular disease, chronic kidney disease, chronic hypertension, and metabolic disease were excluded. Placental tissue was washed with ice-cold saline and soaked in 4% paraformaldehyde, followed by paraffin embedding. PE was diagnosed in accordance with the American College of Obstetrics and Gynaecology criteria.^[14]^ The characteristics of the cases are listed in Supplementary table 1.

### Animal Model

Serum in the anticoagulant tube was separated and frozen as aliquots at 80°C until for further use. C57BL/6 wild type mice(purchased from Hunan SJA Laboratory Animal CO., LTD) were kept in a controlled setting (12 h light/12 h dark cycle) of the Central Research Department of Affiliated Hospital of Tongji Medical College Here. We designated the day of vaginal plug appearance was gestational day (E0).

Pregnant mice were randomly assigned (normal_serum group,n=6; PE_serum group, n=6). We administered i.p injection of preeclampsia serum (100μl) or normal pregnancy serum (100μl) per mouse on E10.^[15, 16]^ And on E17.5, the animals were euthanized, fetal weights were recoded, and theplacental units were collected. From E6.5d to E17.5d, blood pressure was measured every two days with tail-cuff plethysmography (BP-2000 Visitech system, Austin, TX, USA) was used to measure

### Paraffin section, dewaxing, staining and brightfield imaging for Spatial transcriptomics

The paraffin tissue was ice bathed for 30 minutes, and then the microtome (Lecia, HistoCore BIOCUT) was used for continuous section (section thickness 5 μm). Slides were dewaxed and stained with H&E. Slides were dehydrated through an ethanol series (100%, 100%, 95%, 95%, 85%, 70%) and cleared in 100% xylene. After staining with eosin, slides were sealed with glycerin. And then brightfield images were taken(G cell, SQS-12P).

### Spatial transcriptomics

Library preparation slides (standard version of the DynaSpatial Probe product with a chip area of 7.5mm by 7.5mm and a spot resolution of 50μm) used were purchased from the DynaSpatial (http://www.dynamic-biosystems.com/content/?349.html). The The DynaSpatial probe program is completed with DynaBlot equipment (a compact benchtop device that transfers transcriptome probes from standard slides to capture slides, allowing you to perform spatial transcriptome analysis from a wider variety of samples). When ready to begin spatial analysis, the slices can be decross-linked and hybridized with transcriptome probes, after which the DynaBlot device helps transfer these transcriptome probes from the slide to the capture region of the capture slide, and then proceed to complete steps such as probe extension and library construction.

### Library preparation of cDNA for sequencing

The steps were performed as earlier described.^[17]^ Finished libraries were diluted to 4 nM and sequenced on the Illumina HiSeq or NextSeq platform using paired-end sequencing. Typically, Sequencing strategy: PE150, double-ended Index; Recommended sequencing depth: 5000reads /spot; General sequencing 60G. When analyzing sequencing data, the 1-8 bases and 39-46 bases of Read1 are Barcode sequences, and the 47-58 bases are UMI sequences. ST sequencing reads were mapped against the human genome (GRCh38), and Ensembl (release 85) transcripts were quantified, as described previously.^[18]^

### Sequencing quality control and bioinformatics analysis

Fragment quality control was performed with Agilent High Sensitivity D5000 Assay. Quality control requirements: segment length is 200-400 bp; The main peak was concentrated around 275 bp. After obtaining the original SequencedReads sequences, the DynaSpatial package DynamicST was used to combine the gene expression information with the location information of the tissue sections to visually demonstrate the spatial distribution of spot types and differential genes on the tissue sections. Subsequently, these differential genes were functionally enriched to identify the functional characteristics of the subpopulations. Differential gene screening was defined as |avg_log2FC|>0.25 and p_val_adj<0.05. Then the differential genes were annotated functionally. Gene annotation contents include: ENTREZID, ENSEMBL, GENETYPE, OMIM, GO, KEGG, Reactome, etc.

## Result

In this study, we performed Spatial transcriptomics on the decidua (nPE = 1, nNormal = 1) and the villus (nPE = 1, nNormal = 1). After quality control and preprocessing, A total of 9832 (nPE = 4778, nNormal = 5045) Spots were retrieved in the decidua, in which the expression of a mean of 3995 and 2221 genes per Spot, 7831, 3585 unique molecular identifiers (UMI) could be detected. And there were 5493 (nPE), 5345 (nNromal) in the villi, about 9567, 12699 UMIs and 4671, 4886 Genens were captured at each spot (Supplementary Figure S1). PCA principal component analysis was first performed, then the 22 principal components with the greatest contribution were selected for cluster analysis. According to the clustering results, Uniform Manifold Approximationand Projection (UMAP) dimensionality reduction algorithm was used to display the distribution of spot in two-dimensional space (Figure 1A,1B). We identified 16 unique clusters, but 17 clusters of PE decidua (Figure 1C). Figure 1C showed the proportion of each cluster in each group. The proportion of cluster 1, cluster 5, cluster 7 and cluster 8 in each group was significantly different. Seurat compared one subpopulation with all other subpopulations to obtain a list of differential genes between spots and other subpopulations. The top10 differential genes (Marker genes) identified in each cluster were displayed by the overall heat map (Figure 1D).

**Figure 1:**
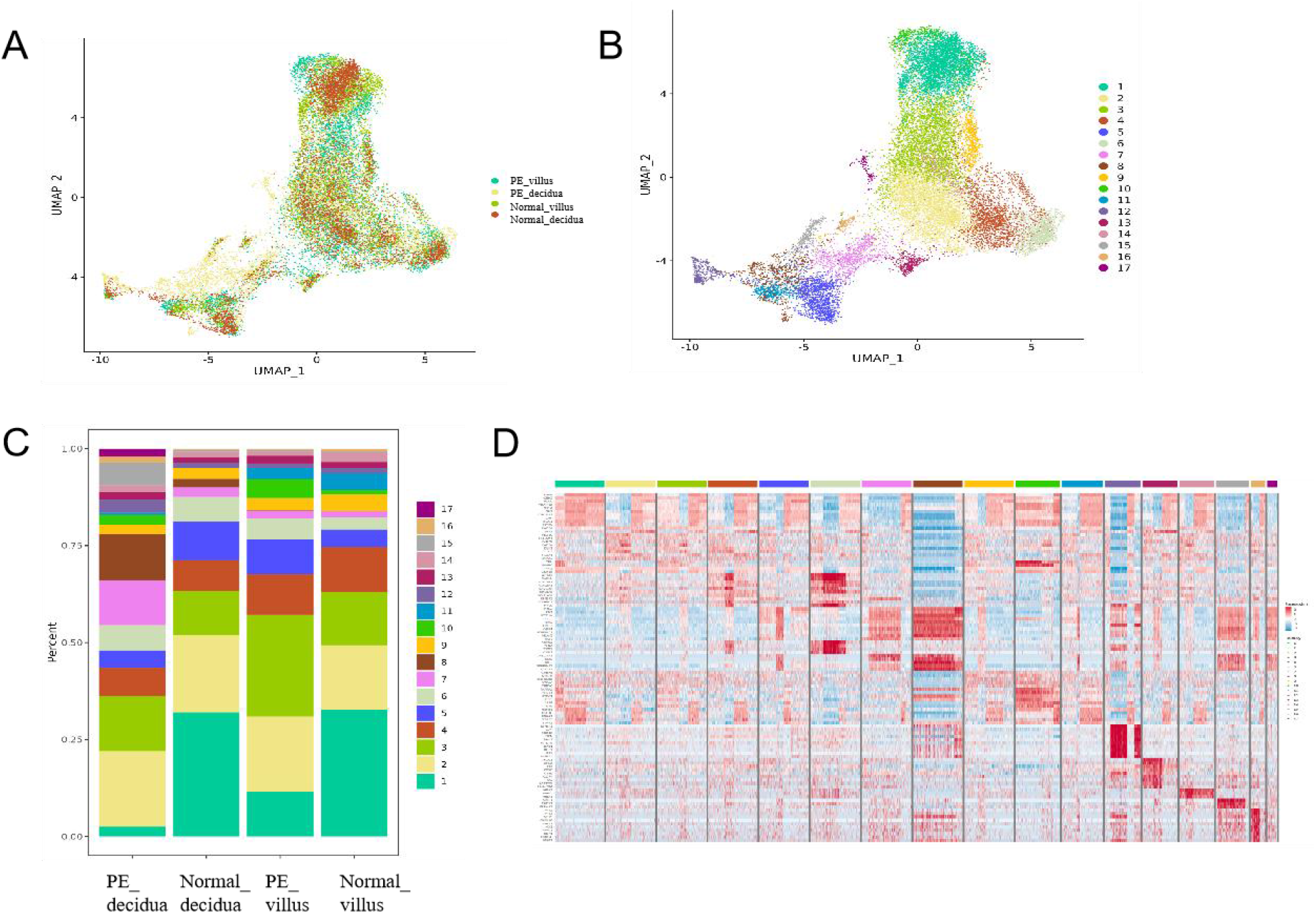
Sample information and cluster dimension reduction. A:Sample dimensionality reduction display UMAP graph; B: Clustering dimensionality reduction display UMAP graph; C: the proportion of each cluster in each group; D: heat map of the top10 differential genes in each cluster.

The UMAP clustering was performed on tissue covered spots from both Normal and PE tissue sections (Figure 2A-2D). Location-specific genes can be detected by the method of markvariogram, which can measure the relationship between gene expression and its spatial location. In Supplementary Figure S2, we show the top 10 genes with spatial differential distribution. In PE and Normal decidua tissues, A total of 1246 differentially expressed genes were screened (Figure 2E). Comparing with the Normal_decidua, AOC1, FSTL3, NOTUM, TAC3, IGFBP1were highly expressed in the PE_decidua. Cell adhesion, cell migration, response to wounding, tissue development, cell motility were mainly involved in the biological process of PE (Figure 2F). And then KEGG and Reactome enrichment analysis were performed on upregulated differential expressed genes (DEG) in PE group. Figure 2G showed the top 20 pathways in KEGG. Focal adhesion, Human papillomavirus infection and ECM-receptor interaction ranked the top 3. The Reactome enrichment analysis showed Extracellular matrix organization, Non-integrin membrane-ECM interactions, Interferon alpha/beta signaling were the top 3 pathways(Figure 2H).

**Figure 2:**
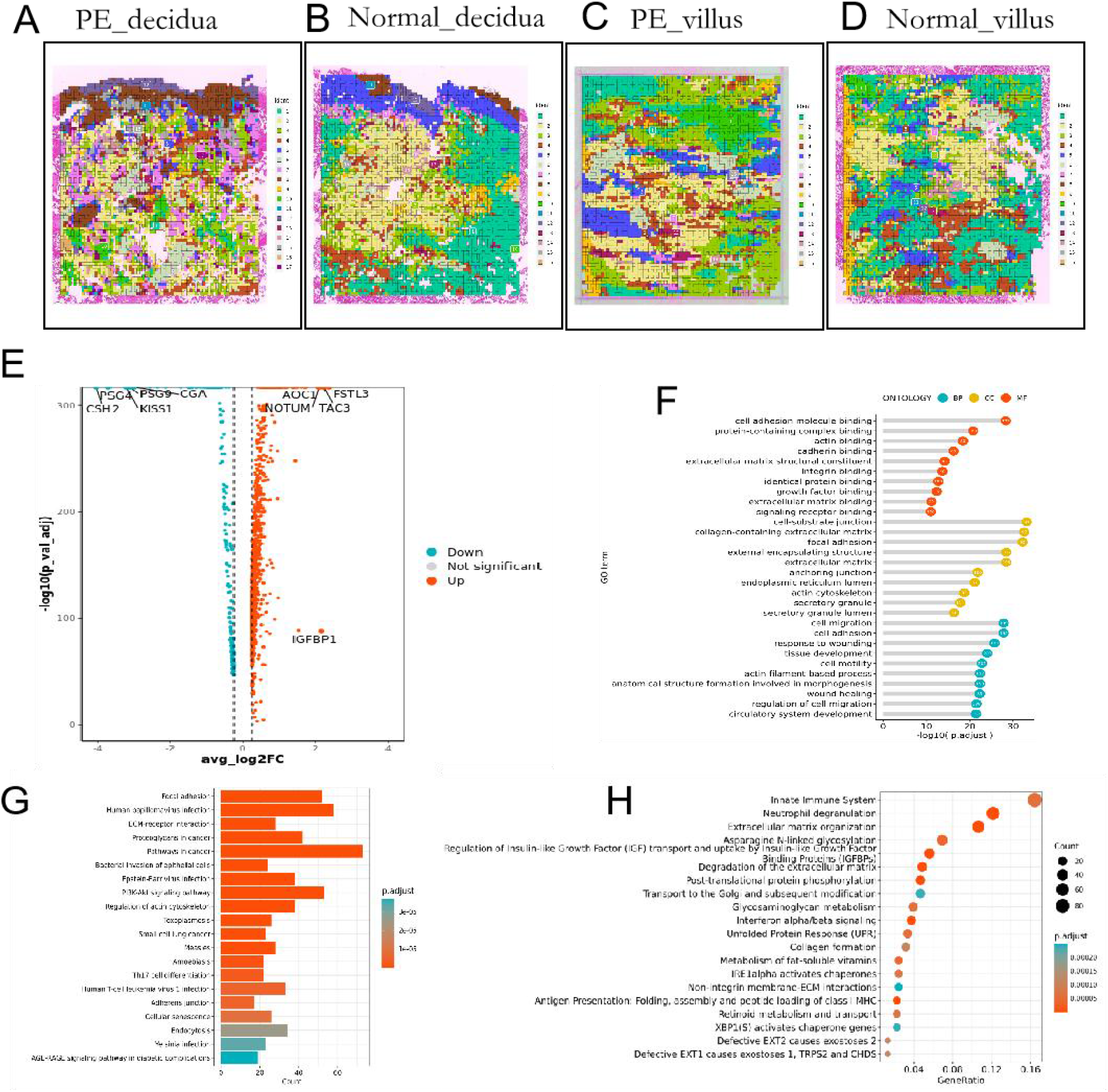
Analysis of differential genes in PE_decidual and Normal_decidual. A-D: Cluster diagram of the four subgroups in spatial transcriptomics; E:differential genes were displayed on volcano map in two group; F: GO analysis of highly expressed genes in PE_decidual; G, H:KEGG(G) and Reactome (H) enrichment analysis of highly expressed genes in PE_decidual.

Cluster 8 is the cytotrophoblast shell, which is the mother-fetal interface junction. The points of cluster 8 were mapped back to the original coordinates in the tissue slices (Figure 3A,3B). The UMAP dimensionality reduction algorithm can effectively project large data sets into two or three dimensions so that similar spots are clustered together. The top10 (sorted by log2FC) highly expressed genes in cluster 8 were specifically labeled (Figure 3C). GO analysis results showed the top 3 term were cell migration, cell motility, and regulation of cell migration (Figure 3D). Figure 3E showed that both FN1 and SERPINE2(mesenchymal marker gene) may play an important role in the 3 terms. Cluster17, which only existed in the PE decidua, had attracted our attention. Supplementary Figure 3A shows the top10 DEG (sorted by log2FC) in cluster17. ClusterProfiler software was used to enrich the GO function and KEGG pathway for upregulated gene sets. GO analysis showed that the top three terms in cluster 17 were tissue development, cell migration, cell motility (Supplementary Figure 3). And FN1 may play an important role in the top 3 pathways (Supplementary Figure 3).

**Figure 3:**
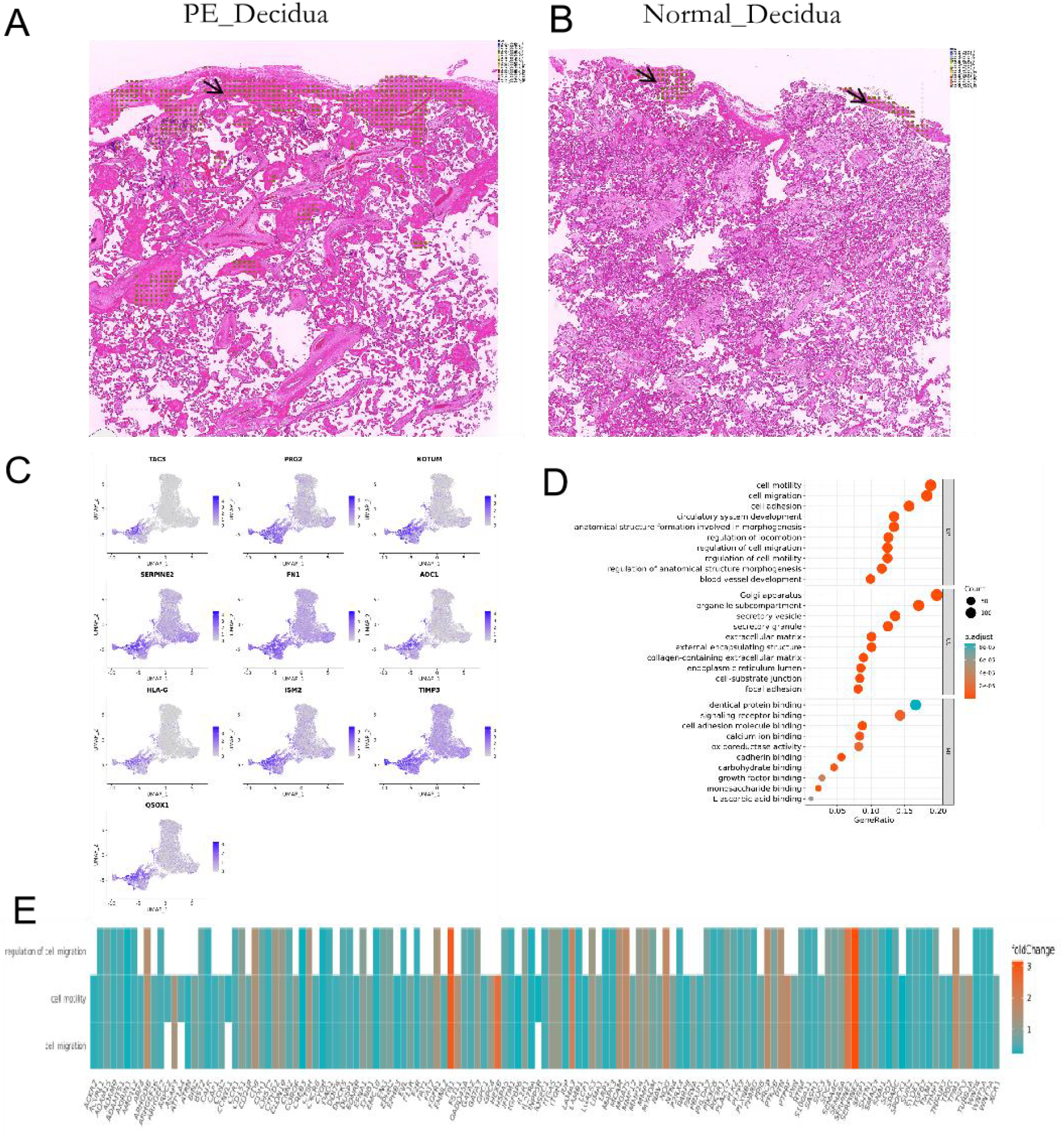
location and gene expression of cluster 8. A-B:Localization of cluster8 in tissue sections; C. UMAP plot of the top 10 genes in cluster 8; D: GO analysis results highly expressed genes in PE_decidual in cluster 8. E: heat map of genes involved in top 3 Terms in cluster 8 of PE_decidual.

Then, we mapped the tissues at the single-cell level combining the cell markers (Supplementary table 2) and spatial transcriptome analysis (Figures 4A,4B). In the cluster 8 of normal decidua were mainly composed of three types of cells: eEVTs, GCs and iEVTs. Surprisingly, the cluster 8 of PE decidua were mainly composed of EVTs) except for GCs and iEVTs. And EVTs in cluster8 of the PE decidua express high levels of TAC3, CDKN1C, IL2RB, SERPINE2, PRG2, HEXB, ITM2B, ADAM19, ISM2, HSPG2 (Figures 4C,4D). GO analysis was performed on the top 10 genes richly expressed in iEVTs, eEVTs and EVTs of PE and normal decidua, and the results showed the main biological process involved in EVTs-PE group was entry into host, movement in host, biological process involved in interaction with host. while in the EVTs-Normal group were negative regulation of plasminogen activation, negative regulation of blood coagulation, negative regulation of hemostasis, negative regulation of coagulation, regulation of phosphatidylinositol 3 −kinase signaling, negative regulation of immune system process(Supplementary Figure 4). GO analysis results also showed the biological processes of the two groups were basically the same in iEVTs(Supplementary Figure 4). HE staining of placental tissue showed a large number of EVTs in the cytotrophoblast shell of PE decidua comparing to the normal decidua (Figure 4E,4F).The immunofluorescence results also confirmed the deposition of EVTs, eEVTs, iEVTs in cytotrophoblastshell of PE group(Figure 4G, Supplementary Figure 5).

**Figure 4:**
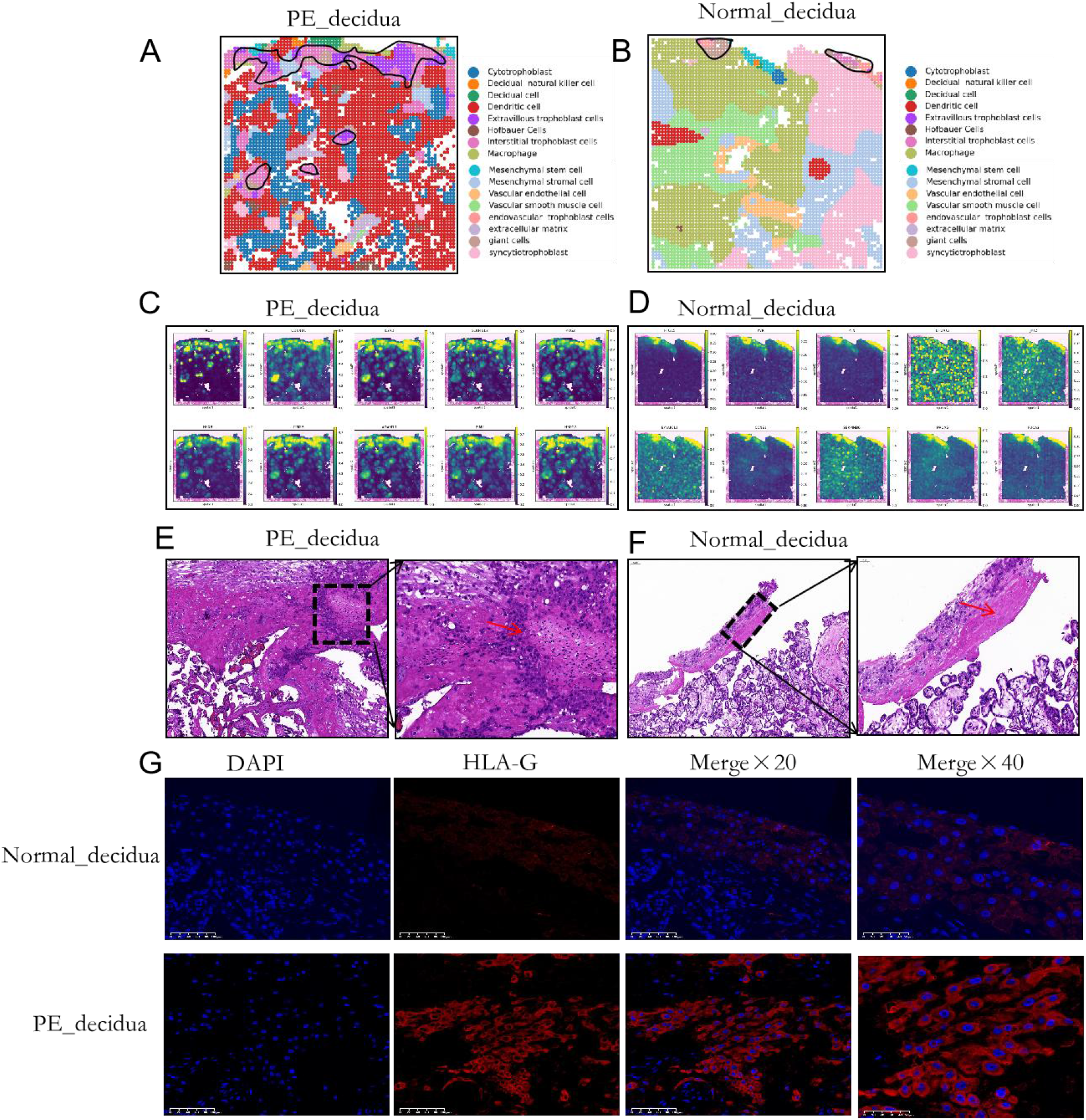
Implantation and defective EVT differentiation in PE_decidua. A-B: the maps of the tissues at the single-cell level; C-D: Spatial maps of the top 10 genes expressed in EVTs in cluster8 of two groups; E-F: HE staining of placental decidua tissue; G: decidua tissue was stained for HLA-G (EVTs marker), Nuclei were stained with DAPI.

Then we performed Monocle2 pseudotime analysis based on the timing of differentiation in the EVTs (Supplementary Figure 6A). Figure 5A showed the top 30 genes expression changes that were differentially expressed over pseudo-time, and the genes were clustered into different clusters based on the similarity of expression. CGA, CSH2, INHBA, PSG3, CSF3R, EBI3, KRT8, SLC40A1were the top 8 genes, and EBI3, KRT8 decreased gradually with the differentiation of EVTs (Figure 5B). Space Focus software (Provided by DynaSpatial) was used to locate the 8 genes at the tissue slice, and we found KRT8 had higher expression in cluster 8 (Figure 5C). Compared with cluster 8 in Normal_decidua, KRT8 overexpression was existed in PE_decidua. Curiously, KRT8 was elevated in the PE_decidual tissue but not significantly different in the PE_villus tissue compared to normal group(Figure 5D, Supplementary Figure 6B).

**Figure 5:**
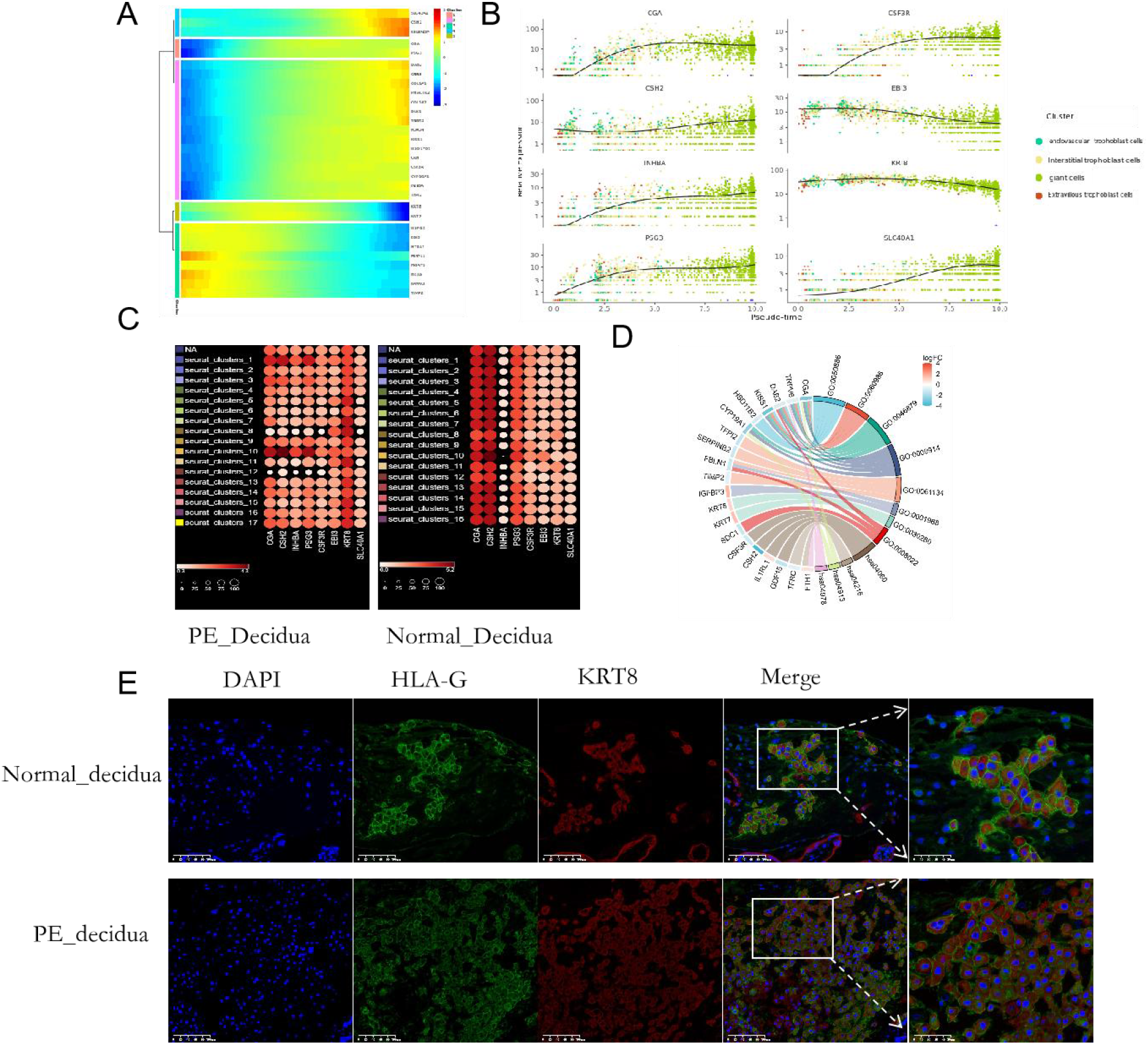
Monocle 2 pseudotime analysis. A: TOP 30 gene expression changes in pseudotime trajectories; B: Changing trends of top 8 genes along differentiation trajectories; C.Distribution and expression heat map of top8 genes in tissue sections D:Chord diagram of differentially expressed genes between PE-decidua and normal_decidua; E: decidua tissue was stained for HLA-G, KRT8, Nuclei were stained with DAPI. Scale bar:100μm

PE-like symptoms models were established by administration of human PE serum (100μl) in pregnant mice (Figure 6A). There were no differences in blood pressure between the two groups before PE serum injection. However, after PE serum injection, both systolic and diastolic blood pressure of the PE_serum group increased significantly (Figure 6B,6C). Figure 6D showed the localization of KRT8 in the placenta of mice in the PE_serum group and Normal_serum group.

**Figure 6:**
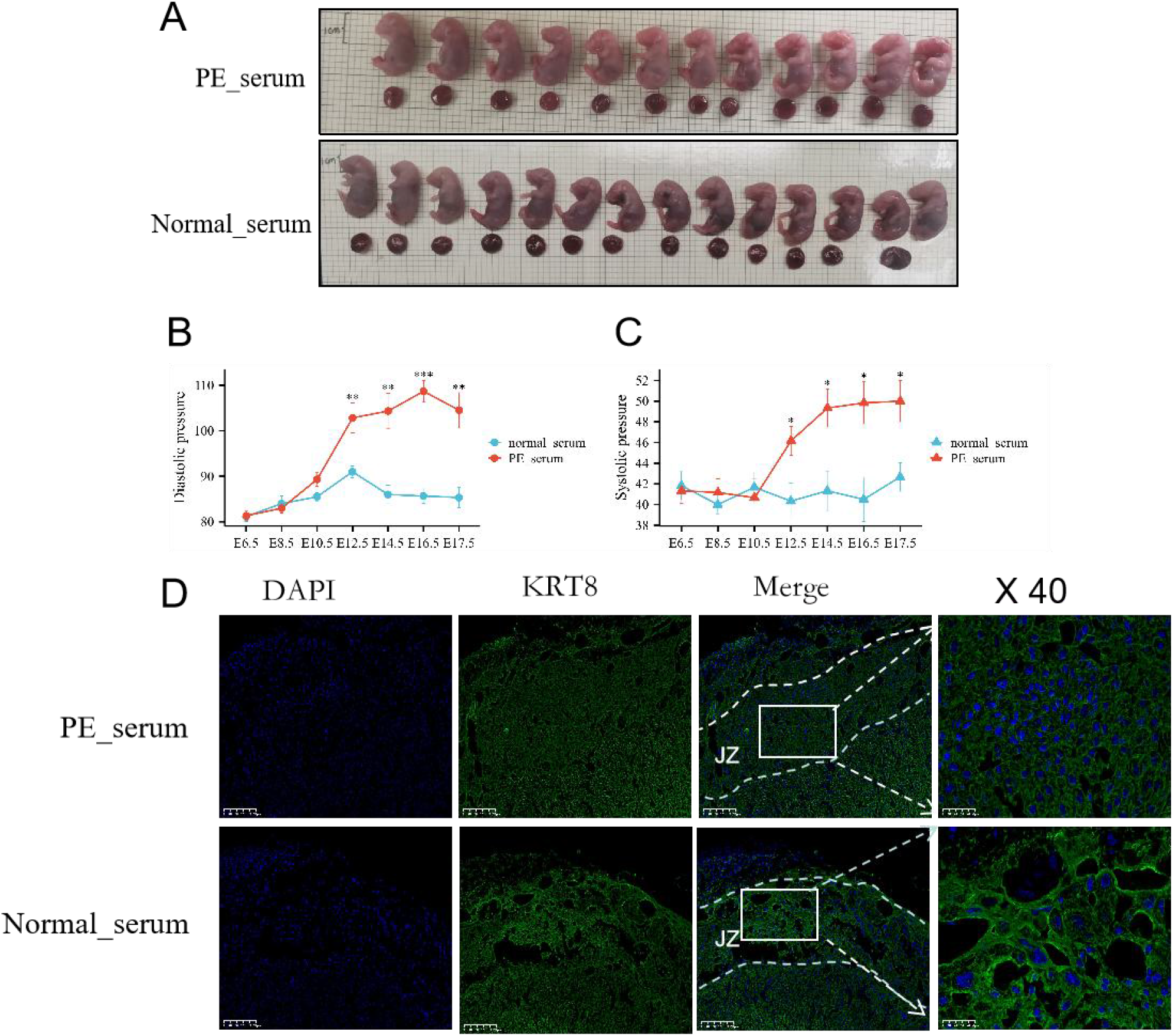
KRT8 in mice with PE_serum induced PE-like symptoms models. A:Representative images of the fetuses in the PE_serum group and the Normal_serum. Scale bar: 1 cm. B,C:Systolic blood pressure(B),diastolic blood pressure(C) of pregnant mice in the PE_serum group and the Normal_serum group.(D) Mouse placenta was stained for KRT8, Nuclei were stained with DAPI. Scale bar:200μm°JZ:Junctional zone.

## Discussion

Preeclampsia, one of the leading causes of premature birth and maternal-infant death, remains a major unaddressed public health problem. Although it is widely believed defects in trophoblast invasion or arterial transformation established during early pregnancy may lead to preeclampsia,^[19]^ the exact mechanism still needs to be further studied.

In this study, we collected preeclamptic and normal placental tissues for ST to further elucidate the mechanism of PE. 17 clusters were identified, including six types of trophoblast cells : SCTs, STBs, EVTs, iEVTs, eEVTs and GC. The differentiation and development of CTBs and EVTs are two crucial processes, which are tightly regulated. And any abnormal regulation can lead to the occurrence of pregnancy-related diseases such as preeclampsia.^[20, 21]^ As we know, EVTs originates from the cytotrophoblast shell (Cluster 8), where is the mother-fetal interface junction. Therefore, we focused on the cluster 8. Combined with cell marker, we found a large amount of EVTs were accumulated in the cluster 8 of PE decidua, and this result was also confirmed in HE stained sections. Monocle2 pseudotime analysis revealed that KRT8 may be the key molecule for EVTs aggregation in cluster8 of PE group, which increased significantly in the decidua tissue of PE, but not in the villi tissue.

KRT8 known as cytokeratin-8 (CK-8) or keratin-8 (K8) is a keratin protein that is encoded by the KRT8 gene in human.^[22]^ KRT8 is essential for the development of mouse embryos.^[23]^ The deletion of the KRT8 gene in the placenta can lead to the loss of sponge trophoblast and labyrinth trophoblast cells, and the proliferation of trophoblast giant cells, thus affecting the function of the placenta, and ultimately leading to fetal death.^[23]^ Under diverse stress conditions, KRT8 can protects retinal pigment epithelium cells against cell death by interacting with the mitochondria through PLEC (plectin) and facilitate the mitochondrial autophagy.^[25]^ KRT8 has also been reported in other studies.^[26-28]^ Elevated KRT8 can lead to decreased differentiation and increased plasticity of alveolar intermediate cells, which can drive the variation of KRAS, and further promote the occurrence of lung adenocarcinoma.^[29]^ In our study, we found KRT8 was over-expressed in the cytotrophoblast shell of PE, but the specific mechanism remains to be further investigated. We will explore the mystery of KRT8’s increase and the possible mechanisms of the accumulation of EVTs.

In summary, we found over-expressed KRT8 in the cytotrophoblast shell led to the accumulation of EVTs, and promoted the occurrence of PE.

## Data Availability

The datasets used and analyzed during the current study are available from the corresponding author on reasonable request

## Declarations

### Ethics approval and consent to participate

The study was approved by the Ethics committee of Maternal and Child Health Hospital of Hubei Province (2021XM049).

## Competing interests

The authors declare that they have no competing interests.

## Funding

Fund of Hubei Province Chutian Talent Program Fund of Major science and technology projects in Hubei Province(No.2022AC005) Fund of Hubei Provincial Department of Science and Technology (No. 2022CFB561)

## Authors’ contributions

YZ and MMC contributed to the protocol design. JP, MHS and QWD collected and analyzed data. MMC drafted the manuscript, XHY, HY, XYC contributed to the interpretation of results. JBW, CYL proofread and commented on the manuscript.KZ revised the final version and are guarantors of this manuscript. All authors made substantial contributions to the paper and read and approved the final manuscript.

## Consent for publication

This manuscript describes original work and is not under consideration by any other journal. All authors approved the manuscript and this submission.

## Acknowledgements

Not applicable.

## Notes

### Competing Interest Statement

The authors have declared no competing interest.

### Clinical Trial

not applicable

### Funding Statement

Fund of Hubei Province Chutian Talent Program; Major science and technology projects in Hubei Province?No.2022AC005?; Hubei Provincial Department of Science and Technology (No. 2022CFB561)

